# Sleep Phenotypes of α-Synucleinopathies and Tauopathies with Parkinsonism

**DOI:** 10.1101/2025.03.13.25323915

**Authors:** Nils Briel, Céline Marti, Esther Werth, Rositsa Poryazova, Philipp Valko, Christian R. Baumann, Heide Baumann-Vogel, Simon J. Schreiner

**Affiliations:** Department of Neurology, University Hospital Zurich, University of Zurich, Switzerland; Clinical Neuroscience Center, University Hospital Zurich, University of Zurich, Zurich, Switzerland; Sleep and Health Zurich (SHZ), University of Zurich, Zurich, Switzerland

**Author notes:** **Corresponding author** Nils Briel, Dr. med., Department of Neurology, University Hospital Zurich, Frauenklinikstrasse 26, 8091 Zurich, Switzerland. equal contribution (Nils Briel, Céline Marti). equal contribution (Christian R. Baumann, Heide Baumann-Vogel, Simon J. Schreiner).

**Keywords:** Parkinson, polysomnography, sleep, α-synuclein, tau, neurodegeneration, RBD/REM sleep behavior disorder

## Abstract

**Introduction:** In neurodegenerative Parkinsonism, biomarkers of α-synucleinopathy (Syn) or tauopathy (Tau) are an unmet need. Rapid eye movement (REM) sleep behavior disorder (RBD) strongly indicates Syn. However, it remains unknown if sleep features other than RBD could reflect underlying neuropathology. Here we assess sleep phenotypes of Syn or Tau in neurodegenerative Parkinsonism and explore their properties as potential biomarkers.

**Methods:** We retrospectively analyzed polysomnography recordings from 198 patients with clinically diagnosed Parkinsonism (20 DLB, 100 PD, 45 MSA, 27 PSP, 6 CBS). We compared sleep features between clinical diagnoses and between Syn (DLB + PD + MSA) and Tau (PSP+CBS) patients. We established linear discriminant analysis-informed parsimonious logistic regression models for differentiating Syn and Tau proteinopathies.

**Results:** Sleep architecture was more disturbed in Tau compared to Syn patients, with less REM and non-REM stage 2 sleep, lower sleep efficiency, and more wake after sleep onset. Stridor was unique to MSA, with a prevalence of 42%. Parsimonious modeling identified sleep features sufficient to differentiate Tau from Syn patients; Diagnostic accuracy was robust with RBD (AUC=0.78) but even higher after adding more polysomnography features (AUC=0.83) and demographic variables to the model (AUC=0.9). The best classification model of Syn vs. Tau is available online for exploration and custom data input at <u>SynTauSleepTool</u>.

**Conclusion:** Distinct sleep phenotypes characterize neurodegenerative Parkinsonism with Syn or Tau. Pending pathological confirmation, our data suggests that neurodegeneration could affect sleep-wake regulatory brain systems in a proteinopathy-dependent manner. Sleep phenotypes hold promise as non-invasive biomarkers of Syn or Tau in Parkinsonism.

**Highlights:**

- Polysomnography in 198 patients with neurodegenerative Parkinsonism.
- Sleep features vary by suspected underlying proteinopathy (Syn or Tau).
- More sleep disturbance in Tau than Syn patients (reduced REM and N2 sleep, more wakefulness).
- Sleep features and demographics accurately differentiate Tau from Syn.
- Sleep phenotypes may have potential as biomarkers of Tau and Syn in Parkinsonism.

**Statement of Significance:** This study highlights how routine polysomnography can reveal distinct sleep phenotypes in neurodegenerative Parkinsonian disorders linked to different underlying pathologies, α-synuclein or tau. By integrating multiple sleep features and demographics instead of relying on isolated well-established sleep biomarkers, such as REM sleep behavior disorder, our approach improves disease classification. These findings underscore the promise of sleep phenotypes as non-invasive biomarkers with potential to guide earlier, more targeted interventions. Future directions include validating these phenotypes in prospectively followed cohorts with confirmed neuropathology.

## 1. Introduction

Sleep-wake disturbances are frequent in patients with neurodegenerative Parkinsonism. While insomnia, or excessive daytime sleepiness are common symptoms in most types of Parkinsonism, other sleep disturbances are rather specific for some types of Parkinsonism, especially rapid eye movement (REM) sleep behavior disorder (RBD). Our and others’ prior studies noted that RBD is highly prevalent in α-synucleinopathies (Syn), including Parkinson’s disease (PD), dementia with Lewy bodies (DLB), and multiple system atrophy (MSA)(Baumann-Vogel et al., 2020). However, RBD is considerably less common in Parkinsonism linked with 4-R tauopathy (Tau), such as progressive supranuclear palsy (PSP) or corticobasal syndrome (CBS)(Boeve et al., 2013). Therefore, RBD is now conceptualized as a biomarker of Syn (Boeve et al., 2013; Cesari et al., 2022; Miglis et al., 2021). Diagnostic guidelines even include RBD in the core criteria for DLB (McKeith et al., 2017). Moreover, RBD along with nocturnal inspiratory stridor may strongly corroborate the diagnosis of MSA(Cortelli et al., 2019). However, focusing on RBD as a biomarker has several limitations. First, the prevalence of RBD increases with age and disease duration, and only a subgroup of Syn patients presents with RBD as a prodromal symptom, limiting the diagnostic value in younger and “brain first” patients (i.e., pathophysiology initiated in the brain rather than the gastrointestinal tract) (Baumann-Vogel et al., 2020; Horsager et al., 2020; Sixel-Doring et al., 2023). Second, common medications, such as antidepressants, can induce RBD (Cortelli et al., 2019). Third, in some patients, loss of REM sleep, presumably due to disease, could render the polysomnography (PSG)-based confirmation of RBD impossible (Bugalho et al., 2021; Cesari et al., 2022).

Sleep architecture refers to the composition and amounts of nocturnal sleep stages, defined by cycling of non-REM (NREM), REM, and wake stages. A complex, finely tuned interplay of brain networks involving cortical, thalamocortical, hypothalamic, limbic, and brainstem regions, regulates sleep architecture. In contrast, RBD is presumably caused by rather focal damage to the lower brainstem resulting in disturbance of related networks and thus incomplete suppression of muscle tone and presentation of parasomnia during REM sleep (Cortelli et al., 2019). Accordingly, RBD is nested in sleep architecture and dependent on the appearance of REM sleep (Bugalho et al., 2021; Cesari et al., 2022). Loss of REM sleep, on the other hand, belongs to the typical changes in sleep architecture in aging and several neurodegenerative diseases (Mander et al., 2017; Walsh et al., 2017). This and other alterations in sleep architecture could provide additional insights into neurodegenerative diseases which may affect specific sleep-wake regulatory brain regions and circuitries more commonly than others. In this line, it has been proposed that the accumulation of predominant misfolded proteins in Syn and Tau may contribute to related disturbances in sleep-wake regulation and sleep macrostructure, through disease-specific impacts on cell type susceptibility and sleep system neuroanatomy (Barone and Henchcliffe, 2018; Son et al., 2024). Consequently, it appears plausible that sleep phenotypes, defined by an array of sleep features, including RBD as well as sleep architecture, could act as non-invasive, yet proteinopathy-specific biomarker.

In this retrospective study, we aim to analyze sleep phenotypes based on routine PSG parameters in a large sample of patients with Syn (DLB, PD, MSA) or Tau (PSP, CBS) who underwent overnight PSG. We chose to analyze routine PSG parameters only to facilitate translation of potential findings into practice. We hypothesized that sleep phenotypes, composed of several sleep features, reflect proteinopathy and improve the diagnostic properties of singular markers such as RBD for differentiating Syn and Tau. To support robust data exploration and inspire future clinical applicability, we developed the interactive *SynTauSleepTool* (https://nbneuro.shinyapps.io/SynTauSleepTool/).

## 2. Methods

### 2.1 Cohort selection

We retrospectively assessed PSG recordings from patients with Parkinsonism recorded in the sleep laboratory at the Department of Neurology, University Hospital Zurich, between 2010 and 2019. Patients underwent PSG for diagnostic purposes, also in the absence of suspected sleep disorders. We included all patients with presumed α-synucleinopathy (PD, MSA, DLB), or 4-R tauopathy (PSP, CBS) recorded until July 2019. Diagnoses of Parkinsonian disorders were applied and reviewed according to international research criteria (Armstrong et al., 2013; Gilman et al., 2008; Hoglinger et al., 2017; McKeith et al., 2017; Postuma et al., 2015). Patients with unclear or overlapping diagnoses were excluded, i.e. only patients with at least “probable” diagnosis were considered. Demographical variables (sex, age), disease duration, and levodopa-equivalent dose (LED) were collected. The local ethics committee (Kantonale Ethikkommission Kanton Zuerich; EK approval No. 2014–0127) approved the study.

### 2.2 Polysomnography

All patients underwent a single-night PSG with digital videography (Embla N7000, RemLogic v3.2; Natus Medical Inc., Pleasanton, CA). Expert examiners with long-standing experience in interpreting PSG recordings of Parkinsonism patients (EW, SJS, RP, PV, and CRB) analyzed all recordings according to the scoring manual of the American Academy of Sleep Medicine (AASM)(Iber et al., 2007). We assessed routine PSG parameters, including N2 latency (N2_latency, min), wake (W) and sleep stages (REM, non-REM N1, N2, N3) expressed as percentages of sleep period time (SPT, i.e. time interval between sleep onset, defined by N2 latency, and lights on), total sleep time (TST), wake after sleep onset (WASO, min), sleep efficiency (SE, TST divided by SPT, %), and indices of arousal, apnea-hypopnea (AHI), oxygen desaturation (ODI), and periodic limb movements during sleep (PLMS). The presence of inspiratory nocturnal stridor was registered by audio recording. We diagnosed RBD and REM sleep without atonia (RWA) according to the International Classification of Sleep Disorders (Sateia, 2014). Additionally, the Epworth Sleepiness Scale (ESS) was obtained.

### 2.3 Statistical assessment

All statistical analyses were performed using RStudio 2024.04.1+748 with R 4.4.0. In the first part of the analysis, we assessed sleep features according to suspected proteinopathy (Syn, Tau) and clinical diagnostic labels (DLB, PD, MSA, PSP and CBS). We computed descriptive statistics and baseline group comparisons for demographic and PSG variables. Categorical variables were assessed using Fisher’s exact or Chi^2^ tests, while continuous variables were evaluated using two-sided T-tests or Kruskal-Wallis tests as indicated. We conducted regression analysis with *glm*, implementing each PSG parameter (referred to as ‘sleep feature’ hereafter) as predictor and the “Proteinopathy” label as response, while controlling for “age”, “sex”, “disease duration”, “antidepressants” and “antipsychotics”. Non-normally distributed continuous response variables were log-transformed, and a “Gaussian” model family was utilized. For discrete response variables, a “Poisson” family was employed. Predictor variables were mean-centered at 0 and scaled to −1 and 1 for comparability. Correction for multiple testing was performed using the “False Discovery Rate” (FDR) method. We applied an Analysis of Co-Variance (ANCOVA, type III) for diagnostic labels, while correcting for the same covariates. Tukey’s post-hoc test was performed for the detection of pairwise group differences. We generated radar plots using *ggradar* (https://github.com/ricardo-bion/ggradar) and forest plots using the model input from regression analyses.

In the second part of the analysis, we assessed the potential of PSG parameters for differentiating Tau and Syn. Linear discriminant analysis (LDA) was performed on the scaled and centered data points of two overlapping feature sets, setting the proteinopathy label as target variable. The input consisted of selected sleep features (i.e., ESS, SPT, TST, SE, REM, N1, N2, N3, W, WASO, N2_latency, arousals, PLMS, AHI, ODI, RWA, RBD, stridor) and sleep features plus demographic/clinical variables (i.e., adding sex, age, disease duration, antidepressants, antipsychotics). We trained both models on the same 80-20 train-test set partitioning of observations. Prior group probabilities were included. Performance of LDA class prediction was conducted at the proteinopathy level. Linear discriminant (LD) variable coefficients were depicted as squared spectrum plots guiding LD axes. Based on the established literature and previous ANOVA and LDA analyses we selected parameters for incremental logistic regression models. We set the proteinopathy status as the response variable and three different combinations of PSG and demographic variables as predictors, as follows:

1. *RBD*: Proteinopathy ∼ RBD + RWA
2. *RBD+Sleep*: Proteinopathy ∼ RBD + RWA + REM + SE + W + Arousal_index + N2 + TST
3. *RBD+Sleep+Demo:* Proteinopathy ∼ RBD + RWA + REM + SE + W + Arousal_index + N2 + TST + age + sex + disease duration

Model performance was evaluated using classification metrics, including area under the curve (AUC) of the receiver operator curve, accuracy, sensitivity, and specificity. To address sampling bias and unstable performance statistics, we employed a 500-fold resampling and averaging strategy. We evaluated model performance on each sample corresponding to a 75%-train-test set split, and the reported results are averages of all resampling outcomes. All source code related to these analyses is available at https://github.com/nes-b/SynTauSleep. The *SynTauSleepTool* was developed using the shinyapps platform to facilitate additional data exploration and is accessible for individual data input at https://nbneuro.shinyapps.io/SynTauSleepTool/.

## 3. Results

### 3.1 Sleep Phenotypes Across Proteinopathy and Diagnoses

We included a total of 198 patients with a probable clinical diagnosis of neurodegenerative Parkinsonism, categorized into presumed proteinopathy with Syn (Table 1, N_Syn_=165: N_PD_=100, N_MSA_=45, N_DLB_=20) or Tau (N_Tau_=33; N_PSP_=27, N_CBS_=6). Patients with Syn were younger, had a longer disease duration, and were on higher LED than those with Tau (all p<0.001). Among diagnosis groups, PD patients were the youngest and had both the longest median disease duration, and the highest LED when compared to non-PD subjects. Patients who were on antidepressant or benzodiazepines/benzodiazepine receptor agonist treatment during PSG were excluded, and for antidepressants, a 2-week washout was required. The prescription of antipsychotics was more common in CBS and DLB cases. Sexes were balanced between proteinopathies but differed between clinical diagnoses.

**Table 1.** Demographic and sleep features across proteinopathy and clinical diagnoses. ^1^Median (IQR); N (%). ^2^ Wilcoxon rank sum test; Pearson’s Chi-squared test; Fisher’s exact test. ^3^Kruskal-Wallis rank sum test; Fisher’s exact test. ^4^False discovery rate.^5^ Antidepressant use prior to washout period.

|  | N | Syn<br>N = 165<sup>1</sup> | Tau<br>N = 33<sup>1</sup> | p-<br>value<sup>2</sup> | N | DLB<br>N = 20<sup>1</sup> | PD<br>N = 100<sup>1</sup> | MSA<br>N = 45<sup>1</sup> | PSP<br>N = 27<sup>1</sup> | CBS<br>N = 6<sup>1</sup> | p-<br>value<sup>3</sup> |
| --- | --- | --- | --- | --- | --- | --- | --- | --- | --- | --- | --- |
| **Age (y)** | 198 | 63 (54-70) | 70 (65-74) | <0.001 | 198 | 73 (68-75) | 61 (51-67) | 63 (54-69) | 71 (68-78) | 64 (61-67) | <0.001 |
| **Disease Duration (m)** | 198 | 54 (24-96) | 29 (19-42) | <0.001 | 198 | 36 (23-62) | 77 (36-135) | 36 (21-48) | 24 (16-48) | 30 (25-35) | <0.001 |
| **Sex (M,%)** | 198 | 106 (64%) | 16 (48%) | 0.089 | 198 | 16 (80%) | 68 (68%) | 22 (49%) | 13 (48%) | 3 (50%) | 0.034 |
| **LED (mg/d)** | 198 | 600 (200-1000) | 160 (0-600) | <0.001 | 198 | 100 (0-534) | 800 (480-1100) | 240 (0-600) | 150 (0-390) | 700 (263-950) | <0.001 |
| **ESS** | 198 | 9.0 (5.0-12.0) | 6.0 (3.0-8.0) | 0.011 | 198 | 9.5 (6.0-12.0) | 9.0 (5.0-12.0) | 8.0 (4.0-12.0) | 6.0 (3.0-8.0) | 6.5 (4.5-8.5) | 0.11 |
| **TST (min)** | 198 | 326 (271-364) | 269 (207-311) | <0.001 | 198 | 334 (280-385) | 320 (270-372) | 334 (278-356) | 271 (212-311) | 250 (208-325) | 0.013 |
| **SPT (min)** | 198 | 415 (374-441) | 421 (378-439) | 0.8 | 198 | 399 (361-442) | 414 (372-440) | 419 (392-440) | 421 (373-439) | 416 (388-436) | >0.9 |
| **Latency to N2 (min)** | 197 | 22 (10-42) | 29 (17-47) | 0.2 | 197 | 27 (12-47) | 17 (8-41) | 29 (14-42) | 29 (17-49) | 27 (17-38) | 0.2 |
| **%Sleep Efficiency** | 198 | 80 (66-89) | 62 (49-76) | <0.001 | 198 | 88 (69-93) | 79 (65-90) | 79 (67-85) | 63 (51-75) | 60 (49-79) | <0.001 |
| **% REM** | 198 | 12 (7-18) | 6 (3-9) | <0.001 | 198 | 9 (7-15) | 14 (7-19) | 9 (5-18) | 6 (4-10) | 5 (2-7) | <0.001 |
| **% N1** | 198 | 11 (7-16) | 11 (7-17) | 0.7 | 198 | 8 (6-13) | 10 (7-16) | 11 (8-16) | 10 (7-14) | 17 (11-18) | 0.3 |
| **% N2** | 198 | 39 (30-47) | 28 (23-36) | <0.001 | 198 | 43 (31-59) | 40 (30-48) | 35 (27-45) | 28 (22-35) | 30 (24-38) | 0.001 |
| **% N3** | 198 | 12 (7-20) | 14 (8-18) | 0.7 | 198 | 12 (7-23) | 13 (7-20) | 10 (7-19) | 14 (9-19) | 10 (4-15) | 0.5 |
| **% W** | 198 | 19 (10-31) | 38 (24-51) | <0.001 | 198 | 12 (6-31) | 19 (10-31) | 21 (14-33) | 37 (25-49) | 39 (21-50) | <0.001 |
| **RBD** | 189 | 107 (68%) | 5 (16%) | <0.001 | 189 | 20 (100%) | 47 (51%) | 40 (89%) | 4 (15%) | 1 (17%) | <0.001 |
| **RWA** | 185 | 112 (72%) | 8 (27%) | <0.001 | 185 | 17 (94%) | 58 (62%) | 37 (84%) | 7 (29%) | 1 (17%) | <0.001 |
| **PLMS Index** | 195 | 1 (0-19) | 14 (1-44) | 0.009 | 195 | 1 (0-30) | 0 (0-5) | 8 (0-56) | 14 (1-53) | 17 (0-41) | <0.001 |
| **Arousal Index** | 198 | 9 (5-14) | 16 (9-18) | <0.001 | 198 | 6 (4-9) | 9 (6-14) | 10 (6-18) | 15 (9-18) | 19 (16-29) | 0.002 |
| **Stridor** | 197 | 13 (7.9%) | 0 (0%) | 0.13 | 197 | 0 (0%) | 0 (0%) | 13 (29%) | 0 (0%) | 0 (0%) | <0.001 |
| **AHI** | 197 | 3 (1-11) | 7 (2-15) | 0.081 | 197 | 3 (2-7) | 2 (1-8) | 8 (3-17) | 6 (2-12) | 12 (6-20) | <0.001 |
| **ODI** | 198 | 4 (1-9) | 5 (2-9) | 0.6 | 198 | 7 (4-9) | 2 (1-7) | 8 (2-18) | 4 (1-8) | 7 (5-14) | 0.004 |
| **Antidepressants use<sup>5</sup>** | 198 | 53 (32%) | 7 (21%) | 0.2 | 198 | 10 (50%) | 28 (28%) | 15 (33%) | 6 (22%) | 1 (17%) | 0.3 |
| **Antipsychotics use** | 198 | 9 (5.5%) | 1 (3.0%) | >0.9 | 198 | 3 (15%) | 3 (3.0%) | 3 (6.7%) | 0 (0%) | 1 (17%) | 0.049 |

Tau patients showed more reduced and fragmented sleep as indicated by shorter TST, less REM and N2 sleep, more wakefulness (W, % of SPT) and WASO (min), and a higher arousal index compared to Syn patients (Fig. 1A, C-E). In the more granular comparisons of clinical diagnoses, CBS patients additionally exhibited a more pronounced N3 reduction with concurrent N1 increase (Table 1, Fig. 1 B-E). Furthermore, respiratory indices such as AHI and ODI differed significantly between clinical diagnoses, but less so between proteinopathies.

**Figure 1.**
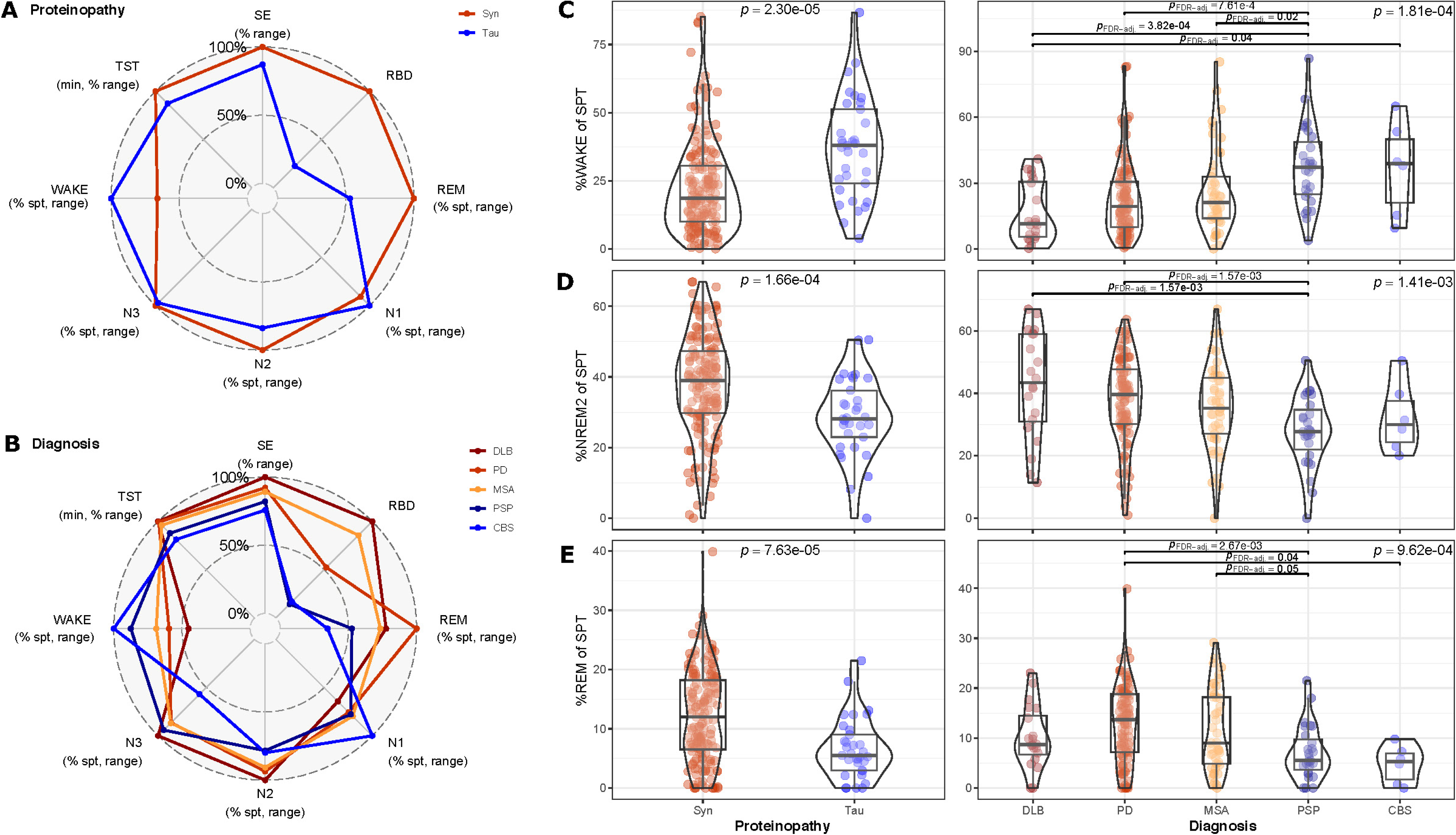
Distinct sleep phenotypes in tau and syn. **A-B** Radar plot with group-wise means of selected sleep features for proteinopathies (A) and clinical diagnosis (B), standardized to 0 and maximum value present. **C-E** Box-violin plot of W (C), N2 (D), and REM (E) (each expressed as % of SPT) comparing proteinopathies (left column, 2-sided Mann-Whitney-U test) and clinical diagnosis (right column; global test statistic: Kruskal-Wallis, post-hoc test statistic: Dunn including false discovery rate, FDR).

To identify independent contributors to the sleep variable-based differential diagnosis of proteinopathies, we implemented multivariable logistic regression. We used normalized and transformed sleep feature data as input and included age, sex, disease duration, and use of antidepressants or neuroleptics as covariates. Thereby, we confirmed independent significant differences between proteinopathies for RBD/RWA, REM, N2, W, and arousal index, among others (Supplementary Table 1, Supplementary Fig. 1A&C).

Next, we compared sleep features across clinical diagnoses while adjusting for covariates as described before (ANCOVA; Tukey’s post-hoc test). The results recapitulated most findings observed at the proteinopathy level, with significant differences in sleep features related to global sleep (TST, sleep efficiency), sleep architecture (REM/non-REM), and qualitative diagnostic features (stridor, RBD) (Supplementary Table 2+3, Supplementary Fig. 1B).

### 3.2 Sleep Phenotypes as Biomarkers of Proteinopathy

Numerous sleep features are inherently interrelated, as they often represent combinations or fractions of one another (for instance, SE is inversely correlated with WASO). To evaluate the combined discriminatory potential of sleep features, we utilized linear discriminant analysis (LDA) on our dataset labelled with clinical diagnoses to gauge proteinopathy differentiation in two competitive approaches. We included either a set of preselected sleep features alone (Fig. 2A, *LDA sleep*, see methods), or a combination of sleep features and demographic and clinical variables (Fig. 2B, *LDA Sleep+Demo*; adding age, sex, disease duration, antipsychotics, and antidepressants use). Both LDA models effectively distinguished between Tau and Syn patients, achieving an overall accuracy of 0.83 (Supplementary Table 4). The balanced accuracy, which takes into account class imbalances, was notably higher in the *LDA Sleep+Demo* vs. the base model (0.66 vs. 0.58). Although expected, these findings emphasize the value of utilizing readily available information such as demographic data for diagnostic considerations.

**Figure 2.**
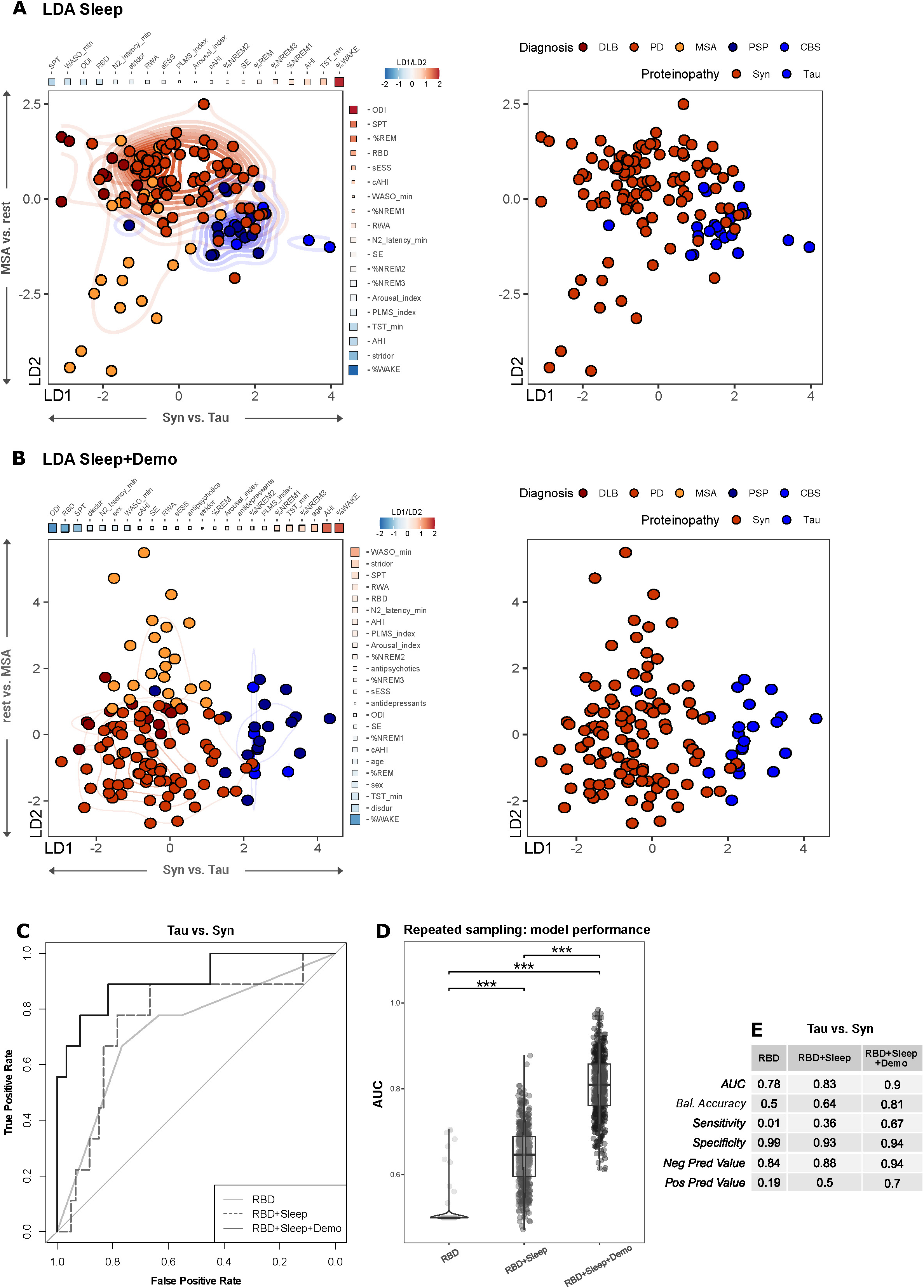
Differentiation of proteinopathies by PSG data. **A** Scatter plot of LD1 and LD2 from LDA on sleep features only; *LDA Sleep*. Color indicates clinical diagnosis (left) and proteinopathy (right). Square spectrum plots guiding the axes indicate LD coefficients (shade). **B** Scatter plot of LD1 and LD2 from LDA on sleep features and demographic data; *LDA Sleep+Demo*. Color indicates clinical diagnosis (left) and proteinopathy (right). Square spectrum plots guiding the axes indicate LD coefficients (shade) **C** ROC of each model with respective AUC from a single train-test set split. **D** Box-violin plot of the average AUC from repeated sampling for each model. ANOVA with Games-Howell post-hoc test, ***P<0.001. **E** Overview of classification performance metrics. Values are means from repeated sampling for each model.

In LDA, the first two LDs per convention capture the most variance between classes, with LD coefficients indicating each variable’s contribution to classification. In the *LDA Sleep* model, LD1 predominantly differentiated Tau from Syn, while LD2 did so for MSA vs. remaining diagnoses (Fig. 2A). The highest absolute LD coefficients were assigned to W, ODI, and SPT. In the *LDA Sleep+Demo* model, LD1 mainly distinguished Tau from Syn, and LD2 did so for MSA vs. remaining diagnoses (Fig. 2B).

To prioritize sleep features, we selected the most consistent parameters from the previous comparisons (i.e., multivariable regression and LDA) and defined three incrementally multivariable models *RBD, RBD+Sleep*, and *RBD+Sleep+Demo* (see methods). We assessed the final classification performance based on averages from 500-fold repeated sampling. The mean classification performance improved from the *RBD* base model (AUC = 0.78, accuracy = 0.5), to *RBD+Sleep* (AUC = 0.83, accuracy = 0.64), and finally to the *RBD+Sleep+Demo* model (AUC = 0.9, accuracy = 0.81) (Fig. 2 C-E).

In summary, these results demonstrate the feasibility of robustly differentiating proteinopathies using PSG-derived sleep features alone. Furthermore, integrating demographic factors substantially refines the interpretive power of sleep evaluations in the differential diagnosis of neurodegenerative Parkinsonism.

## 4. Discussion

Our retrospective cross-sectional analysis of sleep features demonstrates that patients with neurodegenerative Parkinsonism show distinct sleep phenotypes attributable to Tau or Syn pathology and exploitable for diagnostic purposes. These sleep phenotypes extend beyond the presence of RBD and integrate sleep architecture and global sleep parameters into disease-specific sleep signatures (Figure 3). Importantly, selected sleep-related and demographic variables enabled the accurate classification of patients as having Tau or Syn, underscoring the potential of PSG as a non-invasive biomarker. Considering the diagnostic challenges of (atypical) Parkinsonism and the established sleep workup in specialized centers, our data-driven approach to PSG-based disease classification offers valuable insights in a substantial real-world cohort of a pertinent patient population.

**Figure 3.**
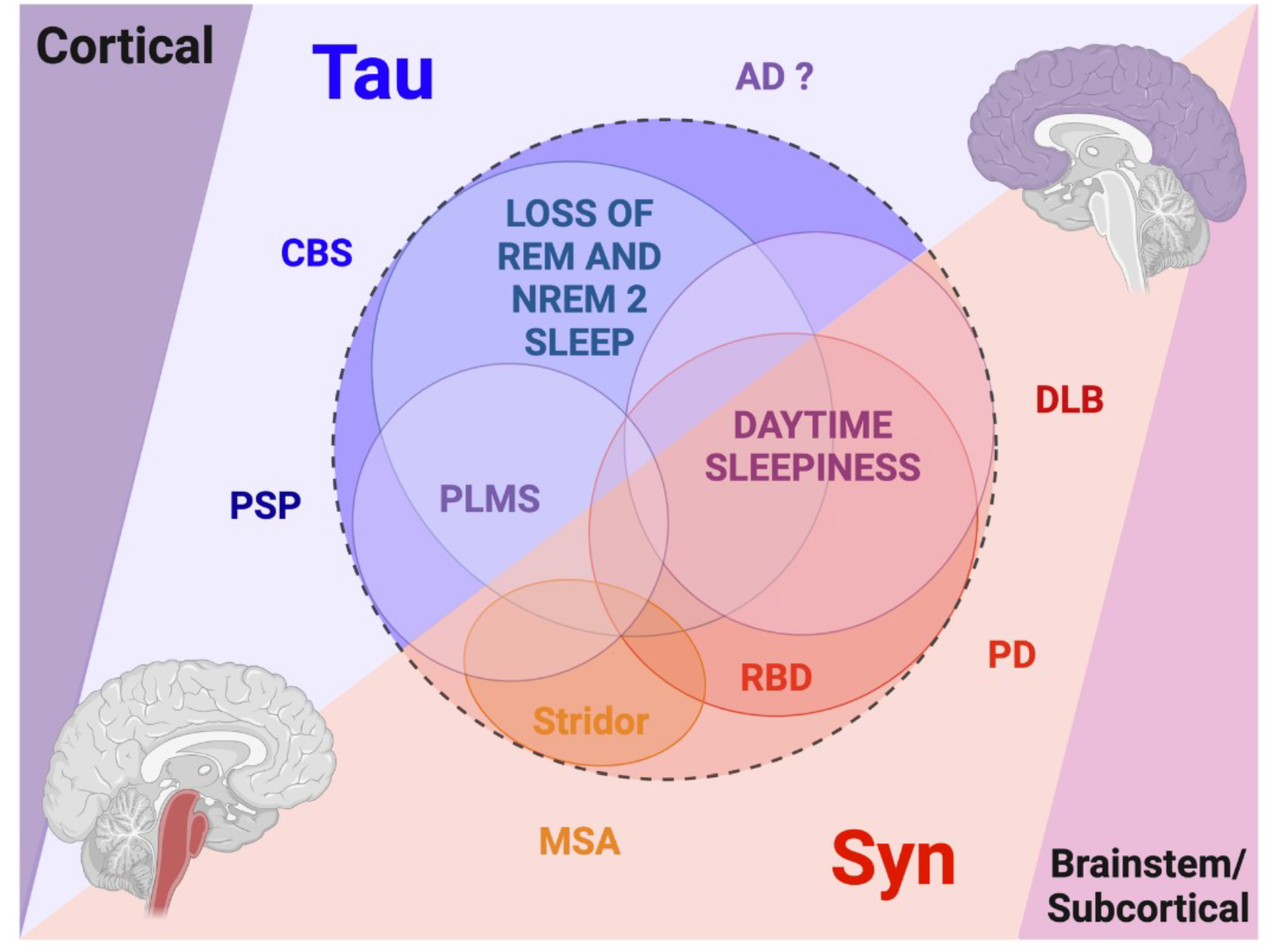
Conceptional framework of sleep phenotypes in Parkinsonism with suspected Syn and Tau pathology. Although most sleep features surpass diagnostic boundaries, with the exception of stridor, specific patterns of sleep features suggest that underlying Syn and Tau pathology may be associated with distinct sleep phenotypes in neurodegenerative Parkinsonism.

### 4.1 Sleep Phenotypes of Proteinopathies

Our study provides new evidence indicating that sleep phenotypes in neurodegenerative Parkinsonism could mirror underlying proteinopathy of Syn or Tau. Sleep phenotypes related to proteinopathy integrate established markers, such as RBD, but also a wider array of sleep features, which can be recorded with routine PSG – a diagnostic requirement for valid RBD diagnosis. As such, our findings corroborate the link between RBD and Syn (Barone and Henchcliffe, 2018; Baumann-Vogel et al., 2020; McKeith et al., 2017), but extends the existing literature by pointing out differences in sleep architecture between patients with presumed Syn or Tau. Specifically, we show that sleep loss and fragmentation is more severe in Tau compared to Syn patients, with reduced SE and TST, more wake, and less N2 and REM sleep. This observation extends previous comparisons of the most common Parkinsonian disorders with Syn or Tau, namely PD and PSP (Sixel-Doring et al., 2009).

Aging is associated with decreased SE, loss of REM and a shift from N3 to N2 and N1 sleep (Sixel-Doring et al., 2009). Additionally, medications such as antidepressants and antipsychotics can alter sleep macrostructure. However, our covariate-adjusted analyses confirm medication-*in*dependent differences in sleep features. In Parkinsonian disorders, adequate dopaminergic substitution can improve sleep fragmentation by reducing motor symptoms (Abbott and Videnovic, 2014). Thus, the higher LED in the Syn group might mediate the preservation of their sleep phenotype as compared to Tau patients. However, we also considered LED as indicative of levodopa-responsiveness, which itself is highly diagnostic and extends to clinical examination findings. Since clinical or motor function was not within the scope of this sleep-focused study, we excluded LED from the multivariable analyses.

We also demonstrate that sleep phenotypes can be used for diagnostic purposes. While RBD is an established biomarker of Syn, our data show that including additional sleep features from PSG recordings improves the diagnostic accuracy to differentiate between Syn or Tau among patients with neurodegenerative Parkinsonism. Using sleep *signatures* as opposed to singular markers (e.g. RBD) is more likely to capture the variability in sleep features, which is probably caused by, and therefore indicative of, disease- and proteinopathy-specific processes. Furthermore, since single or multiple missing data is not uncommon in real-world measurements, composite markers can provide more robustness to interpretation.

Finally, to facilitate broader adoption and inspire practical application of our findings, we developed the interactive *SynTauSleepTool* (https://nbneuro.shinyapps.io/SynTauSleepTool/). This freely-available tool assigns probabilities for proteinopathy and diagnoses from input data, allowing researchers and clinicians to further refine diagnostic insights based on our best-performing classification model for proteinopathy prediction from sleep phenotypes.

### 4.2 Sleep phenotypes and neuroanatomical susceptibility

These findings suggest that the interaction between sleep and neurodegeneration could differ depending on underlying Syn or Tau pathology. In the present data, the divergence in sleep-wake patterns was notably more pronounced when analyzing presumed proteinopathies rather than clinical diagnoses alone. Although this could merely be an effect of group size, findings suggest a distinct neuroanatomical susceptibility within the networks regulating sleep-wake cycles, as previously discussed (Eser et al., 2018; Son et al., 2024; Walsh et al., 2017). In the context of Tau, structures such as the ponto-mesencephalic nuclei, including the *Locus coeruleus* which favors non-REM over REM sleep (Osorio-Forero et al., 2022), and the REM sleep-promoting tegmental nuclei, are consistently impacted by tau protein accumulation and neurodegeneration (Eser et al., 2018). Assuming these pathological alterations are sufficient to disrupt network functions, the observed shift from reduced N2 (to a lesser extent N3), and REM sleep to increased wake in Tau patients seems plausible. Conversely, in Syn, the dorsal mesencephalic-ponto-medullary network appears particularly vulnerable to α-synuclein inclusions, presumably leading to RWA and dream enactment behaviors characteristic of RBD. The amount of N3 did not significantly differ between Syn and Tau patients and was still within the (highly variable) range of age-matched reference data reported in the literature (Mitterling et al., 2015) This is an interesting observation given that disruption of slow-wave sleep (i.e. N3) might play a role in Syn and the most common tauopathy Alzheimer disease (Ju et al., 2014; Morawska et al., 2021; Schreiner et al., 2019). Future studies could decipher features of N3 across Tau and Syn in greater detail, for example, regional and microstructural aspects of N3, such as slow-wave activity.

### 4.3 Sleep Phenotypes Across Clinical Diagnoses

The distinct sleep phenotypes we observed at the level of proteinopathy integrate sleep phenotypes at the level of diagnoses that define groups of Syn or Tau in our study. When focusing on clinical diagnoses, the prevalence of RBD in PD at 51%, DLB at 100%, and MSA at 89%, as well as in PSP and CBS both at 14%, is consistent with our group’s separate cohort and other studies (Baumann-Vogel et al., 2020). Additionally, the significantly disrupted nocturnal sleep in PSP, characterized by reduced TST and sleep efficiency, as well as diminished N2, N3, and REM sleep compared to controls, aligns with the findings of Walsh et al. (Boeve et al., 2013; Walsh et al., 2017). Sixel-Döring et al. also reported poorer sleep efficiency and N2 sleep in PSP compared to PD patients (Sixel-Doring et al., 2009).

Interestingly, despite the pronounced abnormalities in sleep architecture, Tau patients exhibit less pronounced subjective excessive daytime sleepiness, which may be attributed to impaired reflective-memory capability due to frequent frontal lobe involvement in primary tauopathies as speculated in (Sixel-Doring et al., 2009). An alternative explanation could be a loss of sleep drive as proposed in (Walsh et al., 2017).

While research on sleep in CBS is scarce and often hampered by small sample sizes and diagnostic uncertainty, the evidence suggests increased sleep fragmentation and PLMS (Abbott and Videnovic, 2014). In the present study, sleep was most severely altered in CBS and most preserved in DLB compared to other Parkinsonian syndromes. In contrast, previous studies reported more disrupted sleep in DLB vs. PD patients, though not consistently (Townsend et al., 2023). Our findings corroborate a higher PLMS index in CBS and PSP compared to Syn groups, although the large variance limits its diagnostic value. The pathophysiology of PLMS is thought to be related to reduced dopaminergic activity, but the details remain incompletely comprehended (Bliwise et al., 2012).

From a diagnostic standpoint, while there is some overlap in sleep-wake phenotypes, establishing definitive disease assignments based on single sleep features is impractical, as illustrated by RBD. However, the presence of inspiratory stridor in MSA, which we confirmed, is a distinct yet less sensitive feature. We posit that the co-occurrence of RBD and inspiratory stridor in Parkinsonism is indicative of definitive MSA, when brainstem lesions of other origin are ruled out. Nevertheless, the bidirectional relationship between stridor and MSA diagnosis (i.e., stridor might have contributed to the MSA diagnosis) in our cohort calls for further dedicated research. Our more comprehensive approach, incorporating multiple sleep-related and demographic-clinical variables, holds promise for accurately differentiating diseases and proteinopathies.

### 4.4 Limitations

It is pertinent to acknowledge certain limitations. Importantly, clinical diagnoses often do not align with the definitive neuropathological diagnoses in the spectrum of Parkinsonian disorders, especially for tauopathies (Respondek et al., 2020). The absence of definitive diagnostic labels and the potential co-pathology of Syn and Tau in the same individuals could have influenced the disease boundaries within our cohort. This might also explain the observed 10-33% prevalence of RBD in clinically diagnosed tauopathies across several studies, closely mirroring the rate of misdiagnosed “PSP” cases as actual Syn (Respondek et al., 2020). Even though, a clinicopathologic correlation study of 172 cases provides neuropathological evidence of primary tauopathy in patients who exhibited RBD during their lifetime (Boeve et al., 2013). Despite this potential diagnostic ambiguity, we mitigate the risk of misdiagnosis through comprehensive case-by-case reviews by movement disorders experts.

Moreover, due to the small size of certain subgroups, we did not differentiate between PSP, CBS, or MSA subtypes in our analysis. Neither did we characterize the cohort with fluid or imaging biomarkers of Syn, Tau, or Alzheimer’s pathology. This also limits the effects of presumed proteinopathy on measured sleep features, because diagnostic decision boundaries might be less exact. Alzheimer co-pathology could have additional effects on sleep in Parkinsonism, either as co-pathology in PD and DLB or main pathology in CBS (Gomperts et al., 2008; Respondek et al., 2020). Therefore, future studies should include patients with evidence of Alzheimer pathology and combine PSG studies with fluid or imaging biomarkers.

Importantly, patients in our tertiary referral center underwent PSG for diagnostic purposes, to aid in the differential diagnosis of Parkinsonism. Thus, a strength of our study is that patients are unselected for the suspicion of sleep disorders, such as sleep apnea. Nevertheless, many patients showed long disease duration already by the timing of PSG, which is why the diagnostic value of sleep phenotypes during earlier, eventually even preclinical stages of Parkinsonims remains uncertain and should be tested in future studies.

### 4.5 Conclusion and Outlook

In summary, the present study reveals that distinct sleep composite phenotypes mirror the type of underlying proteinopathy in patients with neurodegenerative Parkinsonism. Sleep is more disrupted and its architecture is more disturbed in Tau compared to Syn patients, while the latter show more RBD. These findings suggest specific vulnerabilities to Syn or Tau pathology within sleep-wake regulatory systems. Sleep phenotypes show promise as non-invasive biomarkers for aiding in the differential diagnosis between Syn and Tau.

## Acknowledgements

We thank all patients included in the study, all of whom provided general consent for using their data for research purposes.

## Disclosures

### Funding

None.

### CRediT authorship contribution statement

**Nils Briel:** Conceptualization, Validation, Formal analysis, Software, Visualization, Writing – major review & editing. **Céline Marti:** Conceptualization, Data curation, Formal analysis, Writing – original draft, Writing – review & editing, Methodology. **Rositsa Poryazova:** Investigation, Methodology, Writing – review & editing. **Philipp Valko:** Investigation, Methodology, Writing – review & editing. **Esther Werth:** Investigation, Methodology, Writing – review & editing. **Christian R. Baumann:** Data curation, Project administration, Resources, Supervision, Funding acquisition, Writing – review & editing. **Heide Baumann-Vogel:** Conceptualization, Data curation, Investigation, Methodology, Project administration, Resources, Supervision, Funding acquisition, Writing – review & editing. **Simon J. Schreiner:** Conceptualization, Data curation, Investigation, Methodology, Project administration, Resources, Supervision, Validation, Visualization, Funding acquisition, Writing – review & editing.

### Competing Interest

**Nils Briel:** Financial disclosure: Received research support from Novartis Foundation for Medical-Biological Research, Association Suisse Romande Intervenant contre les Maladies neuro-Musculaires (co-applicant). Non-financial disclosure: None. **Céline Marti:** Financial disclosure: None. Non-financial disclosure: None. **Rositsa Poryazova:** Financial disclosure: None. Non-financial disclosure: None. **Philipp Valko:** Financial disclosure: None. Non-financial disclosure: None. **Esther Werth:** Financial disclosure: None. Non-financial disclosure: None. **Heide Baumann-Vogel:** Financial disclosure: None. Non-financial disclosure: None. **Christian R. Baumann:** Financial disclosure: Competitive third-party grants from Swiss National Science Foundation, USZ Foundation, UZH Foundation, The LOOP Zurich. Non-restricted grant from AbbVie Pharma. Non-financial disclosure: Shareholder of Tosoo AG (sleep modulation technology). **Simon J. Schreiner**: Financial disclosure: None. Non-financial disclosure: None.

### Data and Code Availability

Data can be made available upon reasonable request to the corresponding authors. All source code is available at https://github.com/nes-b/SynTauSleep/. The *SynTauSleep* tool is available at https://nbneuro.shinyapps.io/SynTauSleepTool/.

## Notes

### Funding Statement

This study did not receive any funding.

### Author Declarations

Ethics committee of the Canton Zurich, Switzerland gave ethical approval for this work

