## Supplementary Fig. for "Sleep Phenotypes of α-Synucleinopathies and Tauopathies with Parkinsonism"

#equal contribution

*equal contribution

### Affiliations

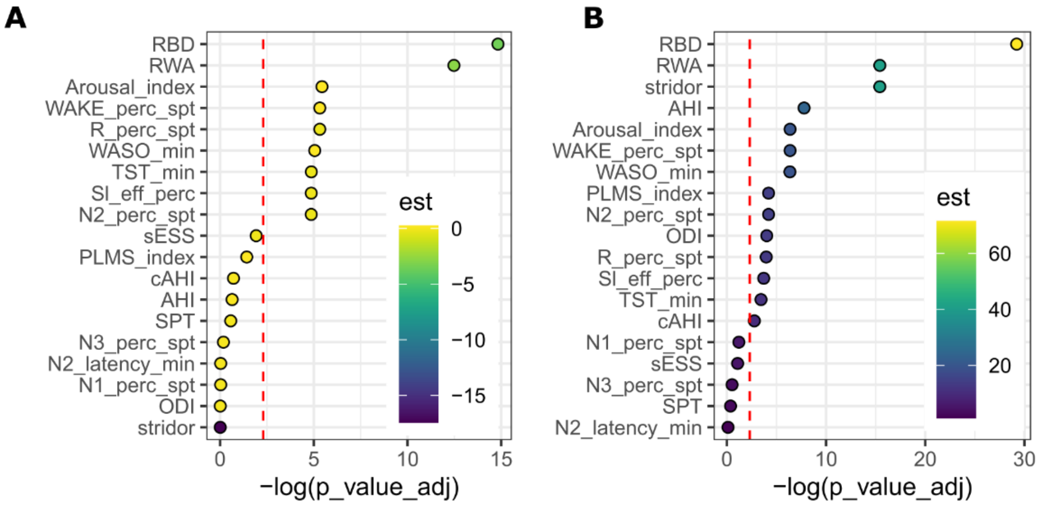


**C**

| Variable | | N | Odds ratio | p |
|---|---|---|---|---|
| **N2_perc_spt** | | | | |
| proteinopathy | Syn | 165 | | |
| | Tau | 33 | 0.78 (0.66, 0.92) | 0.004 |
| sex | F | 76 | | |
| | M | 122 | 1.08 (0.96, 1.22) | 0.201 |
| age | | 198 | 0.90 (0.78, 1.03) | 0.125 |
| disease duration | | 198 | 0.92 (0.81, 1.04) | 0.186 |
| antidepressants | | 138 | | |
| | 1 | 60 | 0.95 (0.84, 1.08) | 0.409 |
| antipsychotics | 0 | 188 | | |
| | 1 | 10 | 1.14 (0.88, 1.48) | 0.335 |
| **RBD** | | | | |
| proteinopathy | Syn | 157 | | |
| | Tau | 32 | 0.03 (0.01, 0.09) | <0.001 |
| sex | F | 73 | | |
| | M | 116 | 0.64 (0.31, 1.31) | 0.232 |
| age | | 189 | 6.87 (2.88, 17.74) | <0.001 |
| disease duration | | 189 | 0.62 (0.31, 1.26) | 0.186 |
| antidepressants | 0 | 135 | | |
| | 1 | 54 | 2.61 (1.16, 6.16) | 0.023 |
| antipsychotics | 0 | 179 | | |
| | 1 | 10 | 1.77 (0.32, 16.73) | 0.561 |
| **WAKE_perc_spt** | | | | |
| proteinopathy | Syn | 165 | | |
| | Tau | 33 | 1.28 (1.11, 1.49) | 0.001 |
| sex | F | 76 | | |
| | M | 122 | 0.92 (0.83, 1.02) | 0.128 |
| age | | 198 | 1.15 (1.02, 1.30) | 0.024 |
| disease duration | | 198 | 1.12 (1.00, 1.25) | 0.046 |
| antidepressants | 0 | 138 | | |
| | 1 | 60 | 1.01 (0.91, 1.13) | 0.820 |
| antipsychotics | 0 | 188 | | |
| | 1 | 10 | 0.80 (0.64, 1.01) | 0.062 |

**Supplementary Figure 1. Polysomnography parameters in adjusted linear models.**

**A** Dot plot showing the negative decadic logarithm of adjusted p-values for covariate-adjusted models assessing differences of PSG parameter across proteinopathies. Color indicates estimates of contribution favoring Syn over Tau. Red line delineates significance threshold of P<0.1 (after Benjamini-Hochberg correction).

**B** Dot plot showing the negative decadic logarithm of adjusted p-values for covariate-adjusted models assessing differences of PSG parameter across clinical diagnoses. Color indicates estimates of contribution favoring MSA over PD. Red line delineates significance threshold of P<0.1 (after Benjamini-Hochberg correction).

**C** Forest plots of multivariable regression analysis for N2, RBD and WAKE. Odds ratios calculated against the reference category (if applicable) and confidence intervals as boxes (middle panel) and numeric (right panel). P values as indicated.

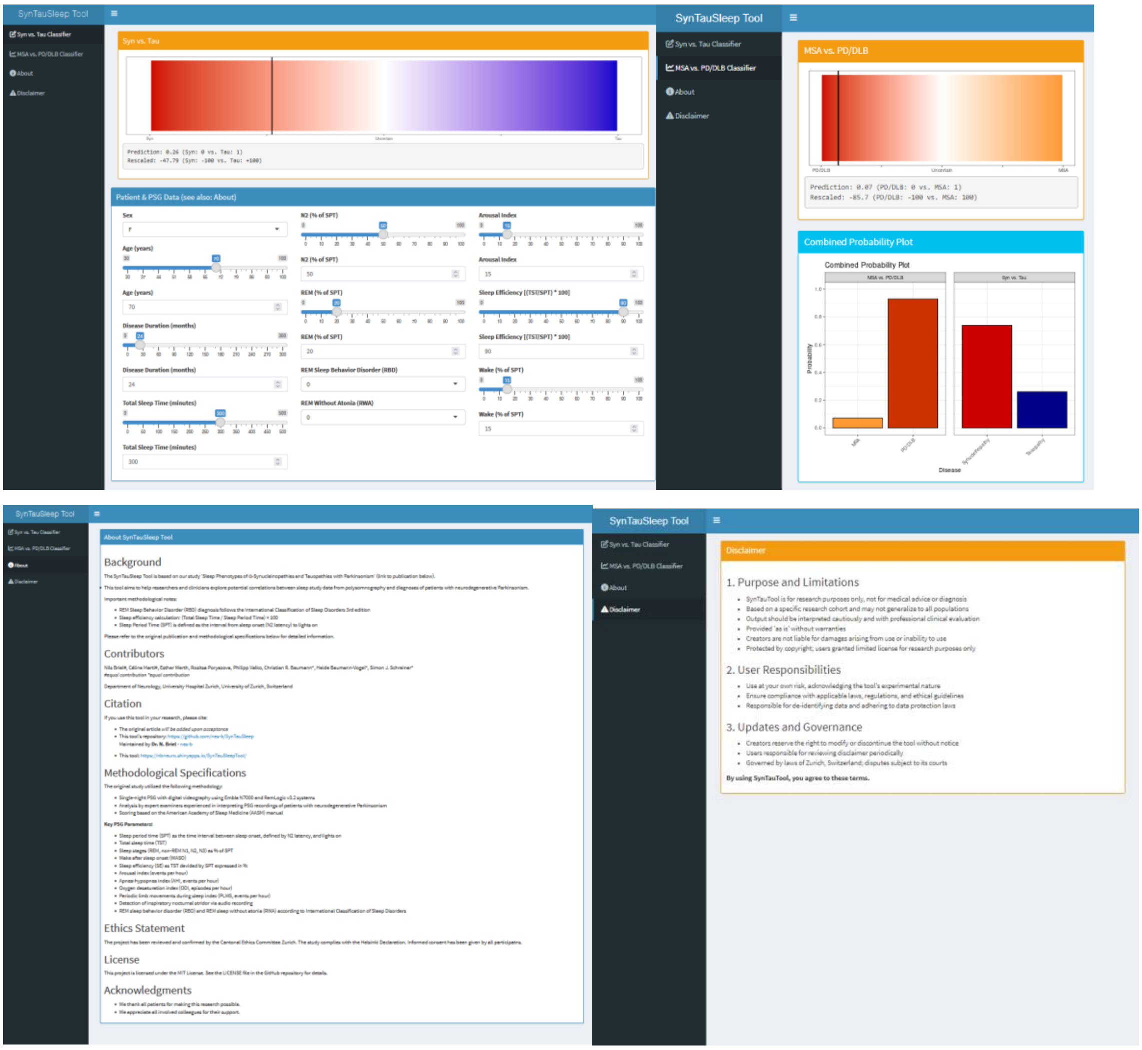


**Supplementary Figure 2. The SynTauSleep Tool.**
The tool displays a combined probability plot for differential diagnosis and includes comprehensive documentation with background information, usage guidelines. The interface is designed to explore sleep-based diagnosis of neurodegenerative Parkinsonism through a user-friendly dashboard.
